# Nonstandard coding of deceased-donor kidney out-of-sequence allocation leads to underrecognition of allocation deviations

**DOI:** 10.1101/2025.02.24.25322786

**Authors:** Emma G. Tucker, Miko E. Yu, Joel T. Adler, David C. Cron, Prateek V. Sahni, Jesse D. Schold, Sumit Mohan, Syed Ali Husain

**Affiliations:** Division of Nephrology, Department of Medicine, Columbia University Medical Center, New York, NY; Columbia University Renal Epidemiology Group, New York, NY; Department of Epidemiology, Mailman School of Public Health, Columbia University, New York, NY; Department of Surgery and Perioperative Care, Dell Medical School, University of Texas at Austin, Austin, TX; Department of Surgery, Massachusetts General Hospital, Boston, MA; Department of Epidemiology, School of Public Health, University of Colorado—Anschutz Medical Campus, Aurora, CO; Department of Surgery, University of Colorado—Anschutz Medical Campus, Aurora, CO

**Keywords:** kidney allocation, organ procurement and transplantation network, organ procurement organization, kidney transplantation, deceased donor

## Abstract

Out-of-sequence (OOS) allocation, the process by which organ procurement organizations (OPOs) can deviate from standard rank lists of potential recipients to expeditiously allocate deceased-donor kidneys, is rising in the U.S. We aimed to determine whether current OPO reporting practices obscure the extent of OOS allocation. Using match-run data for all U.S. deceased-donor kidney transplants from 2021-2023, we defined “miscoded” OOS (mOOS) allocation transplants as those with use of the 799 or 898 OPO- initiated refusal codes (“other, specify”) with free text responses clearly indicating OOS allocation, and compared these to “explicit” OOS (eOOS) allocation, wherein OOS transplants are appropriately coded using refusal codes 861-863. We found that the prevalence of mOOS allocation increased from 2021 (122 transplants) to 2023 (430 transplants) and accounted for 12% of all OOS transplants by 2023. Organs allocated via mOOS had a lower median KDPI than those allocated via eOOS (51% vs 55%, p <0.01). While an increasing number of OPOs used mOOS throughout the study period, the practice remained concentrated overall, with 5 “outlier” OPOs performing 66% of mOOS allocations in 2023. These findings highlight the need for stricter oversight of organ allocation and underscore the responsibility of the OPTN to ensure proper data reporting.

## Introduction

In March 2021, the Organ Procurement and Transplant Network (OPTN) changed deceased-donor kidney allocation by replacing the donation service area (DSA)-based model with a distance-based allocation algorithm commonly referred to as “KAS250.”^1^ This policy was intended to increase geographic equity in transplant but has also increased the operational complexity of organ allocation by 1) increasing the number of “local” transplant centers per organ procurement organization (OPO) tenfold, and 2) increasing the number of offers and administrative workload associated with allocating each organ, which is thought to be contributing to a decrease in organ utilization.^2,3,4,5,6^ This reduction of efficiency in the kidney allocation system temporally coincided with increased scrutiny of OPO performance as measured by, among other things, the successful placement of recovered deceased donor organs.^7^ These policy changes and increased scrutiny were followed by a dramatic rise in “out-of-sequence” (OOS) allocation, the process by which an OPO skips over one or more transplant centers and their candidates to allocate an organ to a predetermined transplant center.^8,9^

The purpose of OOS allocation is ostensibly to avoid accruing cold ischemia time and the associated increased risk of organ discard by expeditiously allocating kidneys that are at a high risk of non-utilization to more “aggressive” transplant centers, rather than following the match run algorithm to identify the highest priority candidate and center willing to accept the organ.^10,11^ OOS allocation could also offer the advantage of reduced administrative burden by limiting the number of centers the OPO must contact during allocation. However, kidneys allocated OOS encompass a broad range of quality, and prior analyses have demonstrated that these organs are more likely to be placed with white, privately insured recipients. This raises concerns about OOS allocation potentially exacerbating various inequities in access to transplant – including the geographic inequities that KAS250 was purportedly meant to solve – by giving certain centers and their patients preferential access to the deceased-donor organ pool.^9,10^ Unlike in the UK, where the allocation of kidneys at high risk of discard can occur through a regulated “Kidney Fast Track” pathway, such determinations in the US are at the discretion of OPOs.^12,13^

To understand the drivers and potential impact of OOS allocation, we must be able to accurately quantify its use. OPOs use the allocation exemption codes 861 (“operational OPO”), 862 (“donor medical urgency”), and 863 (“offer not made due to expedited placement attempt”) to denote OPO-initiated OOS allocation, and these codes have been used to study practice changes in recent analyses.^9,10^ However, a 2024 OPTN analysis also references the code 799 (“Other, specify”) in its definition of OOS transplants.^8^ The 799 code was introduced in December 2021 to replace the identical 898 code.^14^ However, the 799 and 898 codes are intended to be used only when the other bypass reasons available are not appropriate.^15^ The differences in the analytical approaches used to describe OOS organ allocation suggest the need for a careful assessment of how OOS allocation is defined in the potential recipient data to allow for an accurate capture of all OOS placements. We aimed to describe the frequency and nature of 799 and 898 code use from 2021 through 2023 nationally and by each OPO. We hypothesized that use of the 799 code is increasing as OOS allocations are becoming more frequent, that the substitution of the 799 code for the defined codes of 861, 862 and 863 represents a potential change in practice by OPOs, and that this practice is limited to a subset of OPOs.

## Methods

We conducted a retrospective cohort study of U.S. deceased donor kidney transplants allocated between 2021 and 2023 using U.S. transplant registry data. The Columbia University Irving Medical Center Institutional Review Board approved this study. The requirement for informed consent was waived, as this study uses deidentified data from a national transplant registry. This manuscript adheres to Strengthening the Reporting of Observational Studies in Epidemiology (STROBE) guidelines. All research activities were consistent with the principles of the Declaration of Istanbul. All statistical analyses were performed using Stata/MP 17.0 (StataCorp, College Station, TX), and tables and figures were constructed in Stata and Microsoft Excel.

This study used data from the Scientific Registry of Transplant Recipients (SRTR). The SRTR data system includes data on all donor, wait-listed candidates, and transplant recipients in the US, submitted by the members of the Organ Procurement and Transplantation Network (OPTN). The Health Resources and Services Administration (HRSA), U.S. Department of Health and Human Services provides oversight to the activities of the OPTN and SRTR contractors.

### Identifying all deceased-donor kidney transplants

The 2021-2023 Potential Transplant Recipient (PTR) kidney datasets encompass roughly 193 million offers for deceased donor kidneys. Using this file, we identified 57,306 single-organ deceased-donor kidney transplants with a match-run date between January 1, 2021, and December 31, 2023. We excluded transplants that did not have a corresponding entry in the TX_KI file, meaning that there was no record of a corresponding Transplant Recipient Registration (TRR) form (n=1,551). If duplicate donor/recipient pairs appeared in multiple match runs, only the first match run leading to a transplant was kept, and the rest were excluded (n=158). The vast majority (n=124/158) of these duplicates occurred prior to the implementation of KAS250 in March 2021. We excluded kidneys from Puerto Rico’s OPO (n=256) and those with a missing OPO code (n=10) (Supplemental Figure 1).

### Identifying out-of-sequence transplants

We identified a transplant as being “explicit OOS” if the following conditions were all met:

1. The match-run contained one of the following conventional out-of-sequence codes: 861, 862, or 863 AND
2. The OOS code appeared earlier in the match-run than the organ acceptance code.

We identified a transplant as being “miscoded OOS” if:

1. The match-run contained the code 799 (other, specify) or 898 (other, specify), and the accompanying free-text field contained one of the following OOS-equivalent terms: “expedited”, “aggressive”, “open offer”, or “out of sequence” AND
2. The OOS-equivalent code appeared earlier in the match-run than the organ acceptance code AND
3. The match-run did **<u>not</u>** contain one of the following conventional out-of-sequence codes: 861, 862, or 863.

We used Stata to identify free-text fields containing the aforementioned OOS-equivalent terms, then we manually reviewed the remaining free-text fields to identify misspelled entries. Misspelled OOS- equivalent terms (i.e. “agressive”, “expidited”) were reviewed and included in miscoded OOS counts.

### Comparing donor characteristics

We compared donor characteristics of explicit OOS and miscoded OOS organs across all three study years (Table 1). We also compared donor characteristics of explicit and miscoded OOS organs to in-sequence organs (Supplemental Table 1). Donor characteristics included age, gender, race, ethnicity, blood type, KDPI, diabetes, hypertension, peak creatinine, proteinuria, hepatitis C status, cause of death, and donation after circulatory death.

**Table 1:** Donor characteristics of deceased-donor kidneys transplanted via explicit out-of-sequence (eOOS) allocation and miscoded OOS (mOOS) allocation, 2021-2023.

|  | Explicit OOS | Miscoded OOS | p-value |
| --- | --- | --- | --- |
| **Total** | 5,581 (87%) | 854 (13%) |  |
| **Age** | 45 (34-55) | 44 (32-53) | 0.03 |
| **Gender** |  |  |  |
| Female | 2,094 (38%) | 3,476 (62%) | 0.33 |
| Male | 306 (36%) | 547 (64%) |  |
| **Race** |  |  |  |
| Asian | 116 (2%) | 10 (1%) | <0.01 |
| Black | 893 (16%) | 115 (13%) |  |
| White | 4,467 (80%) | 722 (85%) |  |
| Other or multiracial | 94 (2%) | <10 (1%) |  |
| **Ethnicity** |  |  |  |
| Hispanic | 738 (13%) | 88 (10%) | 0.02 |
| Non-Hispanic | 4832 (87%) | 765 (90%) |  |
| **KDPI** |  |  |  |
| Median (IQR) | 55 (33-73) | 51 (30-69) | <0.01 |
| ≥ 85% | 564 (10%) | 71 (8%) | 0.1 |
| **Blood type** |  |  |  |
| A | 1994 (36%) | 340 (40%) | 0.13 |
| B | 678 (12%) | 102 (12%) |  |
| AB | 88 (2%) | 14 (2%) |  |
| O | 2,810 (50%) | 397 (47%) |  |
| **Diabetes** | 654 (12%) | 99 (12%) | 0.99 |
| **Hypertension** | 2,075 (37%) | 316 (37%) | 0.93 |
| **Peak creatinine** | 1.1 (0.7-2.0) | 1.0 (0.7-1.8) | 0.14 |
| **Cause of death** |  |  |  |
| Anoxia | 2,939 (53%) | 443 (52%) | 0.53 |
| Stroke | 1,148 (21%) | 188 (22%) |  |
| Head Trauma | 1,122 (20%) | 178 (21%) |  |
| CNS tumor | 27 (1%) | <10 (0%) |  |
| Other | 334 (6%) | 40 (5%) |  |
| **Donation after circulatory death** | 2,450 (44%) | 344 (40%) | 0.045 |
| **Hepatitis C** | 558 (10%) | 92 (11%) | 0.49 |

3 Abbreviations: OOS, out-of-sequence; KDPI, kidney donor profile index; CNS, central nervous system

Race was included as recorded in the data set, but donors identified as multiracial, Pacific islander, and Native American were categorized as “Other or multiracial.” Donors were classified as hepatitis C positive if they had either a positive hepatitis C antibody test or positive hepatitis C nucleic acid amplification test. We calculated kidney donor profile index (KDPI) from the kidney donor risk index (KDRI) using the scaling factor for the respective year when the match run occurred. Handling of missing data is described in the Supplemental Methods. We compared categorical variables using the chi-squared test and continuous variables using the Kruskal-Wallis test, with a two-sided alpha level of <0.05 as the cutoff for statistical significance.

### Describing trends in miscoded OOS allocation

We calculated the number of explicit and miscoded OOS transplants nationally from 2021-2023, as well as by OPO and by transplant center. We categorized OPOs into three groups based on the number of miscoded OOS allocations performed over the three-year study period. OPOs allocating >40 miscoded OOS kidneys across the study period were classified as “outlier OPOs,” (n=5) while those allocating 15-40 miscoded OOS kidneys were classified as “frequent OPOs” (n=9). All other OPOs were classified as “infrequent OPOs” (n = 42). At the OPO level, we also examined the relationship between the absolute number of mOOS allocations and the proportion of allocations that occurred via the mOOS pathway (Supplemental Figure 2). On January 1, 2023, the Washington Regional Transplant Community (DCTC) OPO merged with the Living Legacy Foundation of Maryland (MDPC) OPO to form the Infinite Legacy (MDPC) OPO.^16^ For this reason, DCTC appears in the “frequent OPO” category in 2021 and 2022, but not 2023, whereas MDPC appears in all three years. Therefore, the total number of OPOs included in Figures 1-2 is 56 in 2021 and 2022 but 55 in 2023.

**Figure 1:**
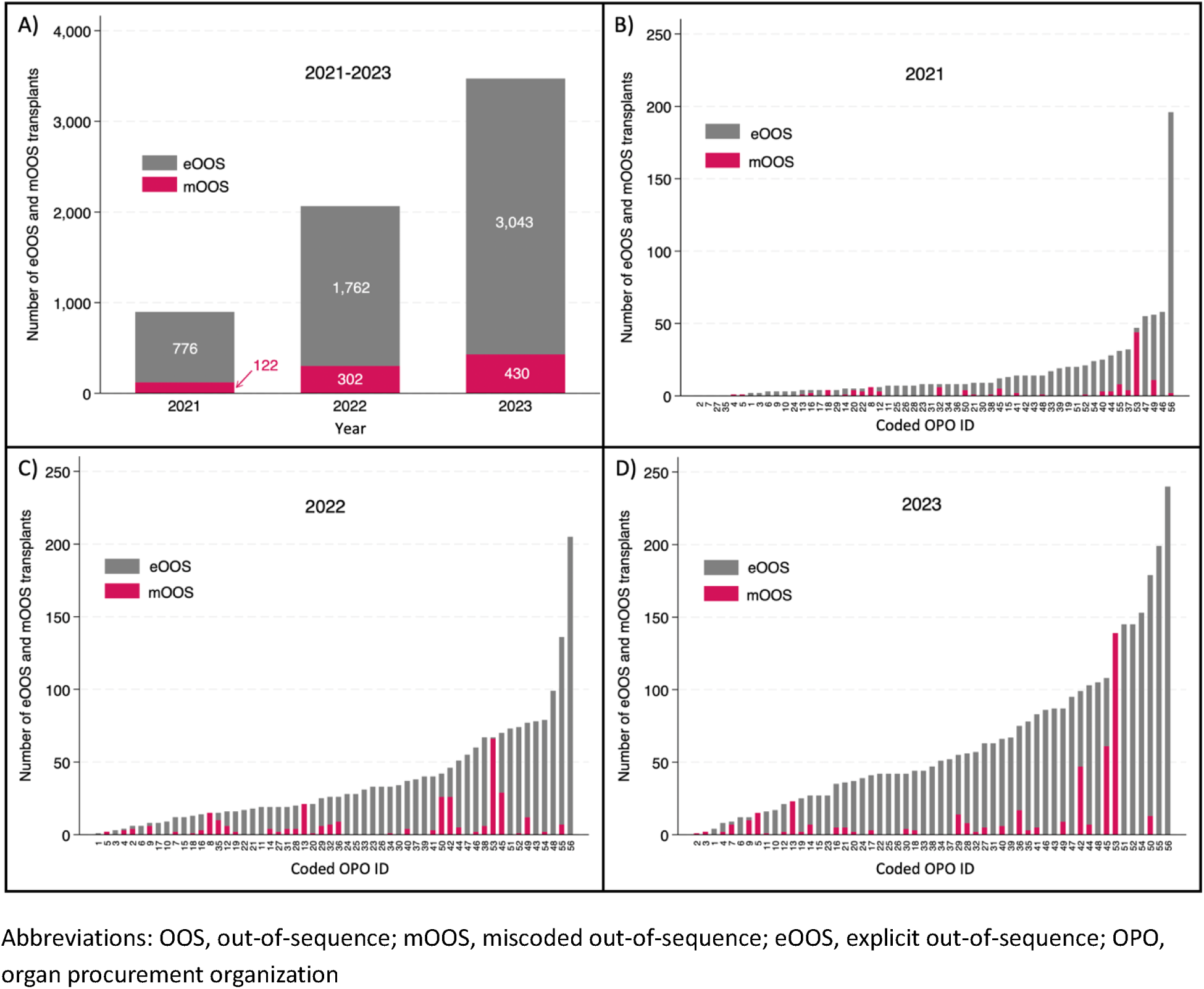
Frequency of explicit and miscoded out-of-sequence allocation at the national level and by organ procurement organization (OPO), by year, 2021-2023. OPOs were assigned numbers 1-56 according to the number of out-of-sequence allocations performed over the study period and maintain the same coded number throughout all panels.

**Figure 2:**
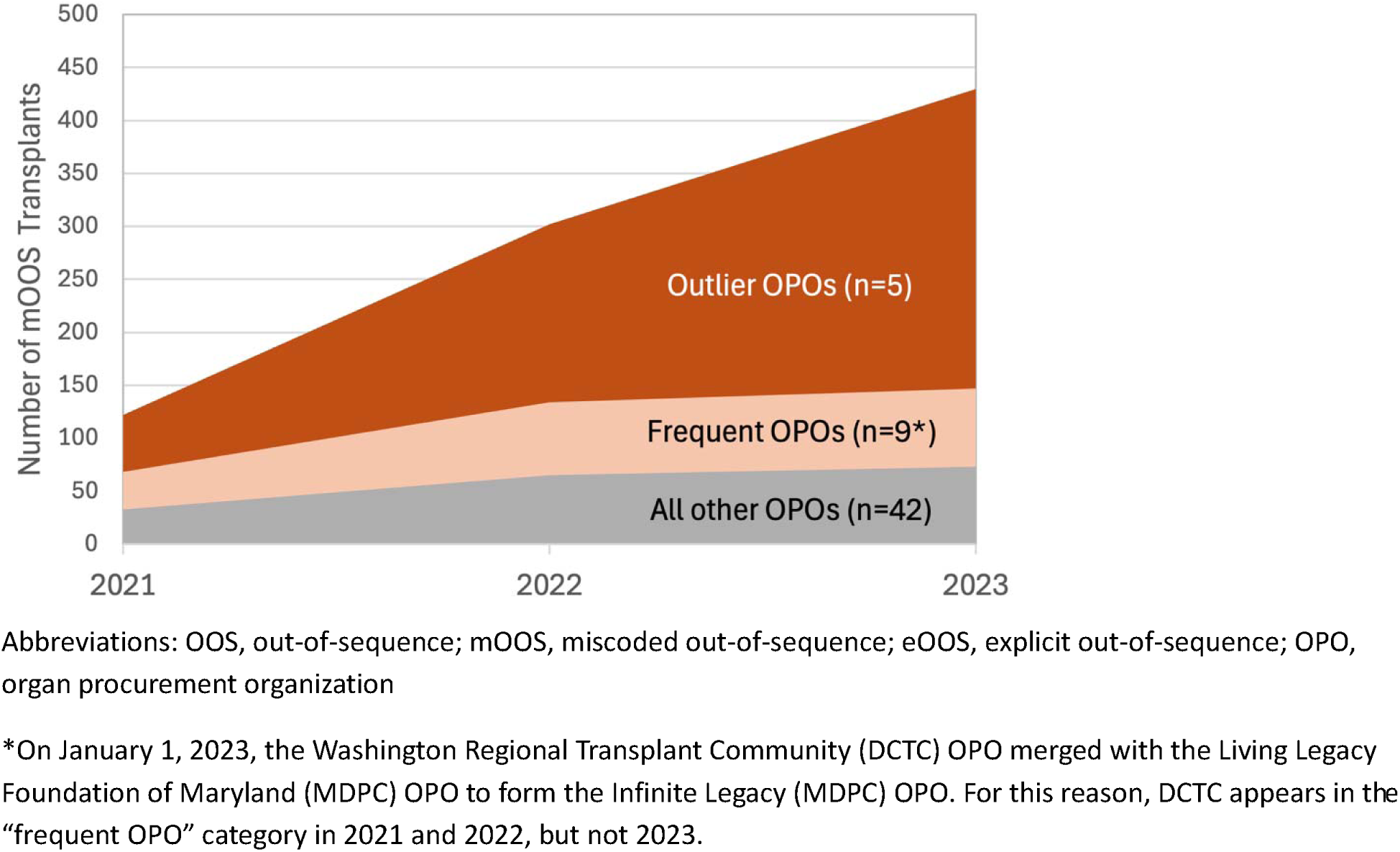
Changes in miscoded out-of-sequence (OOS) allocation from 2021-2023 among outlier organ procurement organizations (OPOs) (>40 miscoded OOS allocations across the study period), frequent OPOs (15-40 miscoded OOS allocations across the study period), and infrequent OPOs (<15 miscoded OOS allocations across the study period)

## Results

Our study cohort included 55,331 single-organ deceased donor kidney transplants, representing 237 transplant centers and 56 OPOs (Supplemental Figure 1). Among these, 5,581 kidneys (10%) were allocated via explicit OOS, meaning their match-runs contained ≥1 OPO-initiated allocation exemption code (861, 862, and 863), whereas another 854 kidneys (13% of the combined total of explicit and miscoded OOS kidneys) had match-runs containing a 799 or 898 free-text entry with one or more of the following OOS-equivalent terms: “expedited,” “aggressive,” “open offer,” or “out-of-sequence” and were deemed miscoded OOS transplants (Table 2). Compared to explicit OOS transplants, miscoded OOS transplants used kidneys from donors who were younger, more likely to be white and non-Hispanic, and had lower KDPI (median 51% vs 55%, p < 0.01) (Table 1).

**Table 2a:** Frequency of out-of-sequence-equivalent terms in the 799 or 898 free text response field of miscoded OOS transplants. Counts indicate the number of match runs containing each term.

| Term | 2021 (n = 122) | 2022 (n = 302) | 2023 (n = 430) | Total (n = 854) |
| --- | --- | --- | --- | --- |
| Open offer | 32 (26%) | 170 (56%) | 194 (45%) | 396 (46%) |
| Expedited | 84 (69%) | 109 (36%) | 178 (41%) | 371 (43%) |
| Aggressive | 9 (7%) | 44 (15%) | 76 (18%) | 129 (15%) |
| Out of sequence | 3 (2%) | 2 (1%) | 6 (1%) | 11 (1%) |

\*Column percentage exceeds 100% because multiple terms may appear in the same match run

**Table 2b:**
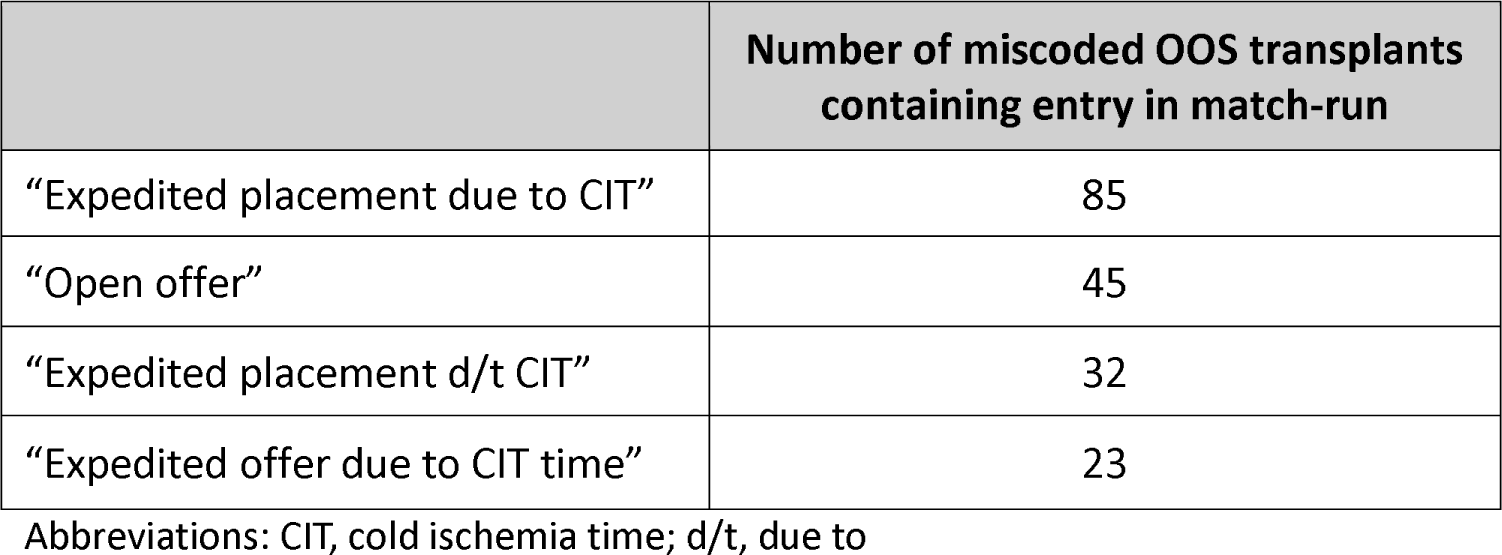
Examples of free text entries used in the match runs of miscoded OOS kidneys.

“Open offer” was the most frequently used OOS-equivalent term, appearing in 46% (n=396) of all 854 miscoded OOS transplant match runs (Table 2). “Expedited” and “aggressive” were the next most frequently appearing terms and were present in 43% (n=371/854) and 15% (n=129/854) of miscoded OOS match-runs, respectively. “Out-of-sequence” was present in 1% of miscoded OOS match-runs (n=11/854).

The overall prevalence of explicit OOS and miscoded OOS allocation increased from 898 kidneys in 2021 to 2,064 kidneys in 2022 – a more than 2-fold increase. This was followed by an additional 68% increase from 2022 to 2023, resulting in a nearly 4-fold increase in just 2 years (898 to 3,473 kidneys) (Figure 1A). The number of miscoded OOS organs transplanted increased nearly 150%, from 122 kidneys in 2021 to 302 kidneys in 2022 and by another 42% from 2022 to 2023. Further, while 25 OPOs used miscoded OOS allocation in 2021, this rose to 34 OPOs by 2022 and 31 in 2023 (Figure 1B-D). The proportion of OOS transplants that were miscoded OOS was similar across study years, from 13.6% in 2021 to 14.6% in 2022 to 12.4% of all OOS kidney in 2023.

Five “outlier” OPOs together accounted for the majority (59%) of miscoded OOS allocations throughout the study period (Figure 2). These OPOs performed 44% (54/122 kidneys) of all miscoded OOS allocations in 2021 and 66% (281/430 kidneys) of all miscoded OOS allocations in 2023. In contrast, the 42 OPOs (75% of OPOs) who least frequently used miscoded OOS allocation accounted for only 20% of miscoded OOS allocations. Although only 2% of all deceased donor kidney transplants were allocated using miscoded OOS allocation in 2023, the proportion of transplanted kidneys allocated via miscoded OOS that year ranged from 0% to 29% within individual OPOs. Outlier OPOs were larger than non-outlier OPOs in terms of overall number of kidneys allocated annually (Table 3). There was a strong correlation between the number of mOOS allocations performed by each OPO and the proportion of allocations that used the mOOS pathway (Supplemental Figure 2). At the transplant center level, we also found clustering of OOS transplants at certain centers; across the study period, the top OOS transplant-performing centers accounted for 25% of all OOS transplants, while the top mOOS and eOOS-performing centers accounted for 27% and 26% of all mOOS and eOOS transplants, respectively (Supplemental Figure 3).

**Table 3:** Characteristics of organ procurement organizations (OPOs) according to the number of miscoded out-of-sequence (mOOS) allocations performed. Outlier OPOs are defined as those performing >40 mOOS allocations across the study period, while frequent OPOs performed 15-40 mOOS allocations, and infrequent OPOs performed <15 mOOS allocations across the same period. The median number of kidneys in each category is shown, with the range in parenthesis.

|  | Outlier OPOs (n=5) | Frequent OPOs (n=9*) | Infrequent OPOs (n=42) |
| --- | --- | --- | --- |
|  | **2021** |  |  |
| Number of recovered kidneys | 603 (436-823) | 459 (283-1110) | 382.5 (97-1337) |
| Number of recovered kidneys transplanted | 400 (341-545) | 313 (198-725) | 239 (67-908) |
| Number of recovered kidneys transplanted via eOOS | 12 (4-47) | 8 (1-56) | 7.5 (0-196) |
| Number of recovered kidneys transplanted via mOOS | 4 (0-44) | 3 (0-11) | 0 (0-4) |
|  | **2022** |  |  |
| Number of recovered kidneys | 670 (433-897) | 540 (288-1214) | 427 (106-1309) |
| Number of recovered kidneys transplanted | 436 (340-528) | 390 (198-746) | 260.5 (63-923) |
| Number of recovered kidneys transplanted via eOOS | 46 (21-70) | 26 (2-136) | 20.5 (1-205) |
| Number of recovered kidneys transplanted via mOOS | 26 (21-66) | 7 (2-15) | 0 (0-10) |
|  | **2023** |  |  |
| Number of recovered kidneys | 871 (472-1094) | 663.5 (332-1339) | 485.5 (89-1296) |
| Number of recovered kidneys transplanted | 551 (327-652) | 444.5 (225-779) | 289.5 (59-898) |
| Number of recovered kidneys transplanted via eOOS | 108 (23-179) | 66 (12-199) | 42 (1-240) |
| Number of recovered kidneys transplanted via mOOS | 47 (13-139) | 9.5 (0-17) | 0 (0-8) |

Abbreviations: OPO, organ procurement organization; OOS, out-of-sequence; mOOS, miscoded out-of-sequence
\*On January 1, 2023, the Washington Regional Transplant Community (DCTC) OPO merged with the Living Legacy Foundation of Maryland (MDPC) OPO to form the Infinite Legacy (MDPC) OPO. For this reason, DCTC appears in the "frequent OPO" category in 2021 and 2022, but not 2023

From 2021 to 2023, consistent with the effort to decrease the use of the “other” field, the overall use of the 799 and 898 codes (i.e. those categorized as miscoded OOS allocation plus those not categorized as such) decreased by 85%. In 2021, these codes were present in the match-runs of 6,150 out of 17,399 (35%) transplanted kidneys, versus 903 out of 19,645 (5%) in 2023. However, the proportion of 799/898 codes that were used with OOS-equivalent terms increased over this period, from 2% in 2021 to 48% in 2023.

## Discussion

In this retrospective cohort study of U.S. transplant registry data, we found that the miscoding of out-of-sequence (OOS) kidney allocations has led to systematic undercounting of OOS allocation in the existing literature. The frequency of miscoded OOS (mOOS) allocation – defined as the incorrect use of the “other, specify” codes instead of established OOS codes to denote OOS allocation – increased rapidly following the implementation of KAS250 in 2021; by 2023, mOOS allocation accounted for 2% of all deceased donor kidney transplants and more than 12% of OOS allocations. The finding that OOS allocation is rising faster than previously described – and the fact that it is increasingly difficult to identify and track these allocation deviations – challenges the ability of researchers and regulators to monitor a pathway that was originally intended as a last resort for marginal organs.

Following the implementation of KAS250, which resulted in a marked increase in complexity and inefficiency in kidney allocation, accurately monitoring allocation deviations has become especially important as OPOs increasingly turn to salvage allocation mechanisms to improve organ utilization.^2,3,4,5,6^ There is currently no consistent methodology for identifying OOS allocations. A recently published analysis highlighting the rapid increase in OOS allocation following KAS250 identified OOS allocation as match runs with the codes 861 (“Operational - OPO”), 862 (“Donor Medical Urgency”), and 863 (“Offer not made due to expedited placement attempt”) prior to organ placement.^1^ By accounting for 799 and 898 codes with free text comments strongly suggesting OOS allocation, we identified mOOS allocations that were previously unrecognized, suggesting a large undercounting of allocation deviations in prior studies. However, this may still underestimate the true prevalence of mOOS, as our process relied on a fixed number of predetermined search terms highly indicative of OOS allocation and is therefore unlikely to capture all instances of mOOS allocation. Without a clear, consistent methodology to track allocation deviations, we cannot ensure transparency and integrity within the transplant system. This is especially important given that only 8% of kidneys allocated via mOOS had KDPIs of 85 or greater, challenging the idea that this pathway is being used primarily for marginal organs and emphasizing the need for closer system monitoring.

There are several potential explanations for the rise in miscoded OOS allocation. Miscoded OOS may reflect human error on the part of OPOs during data entry or inadequate training of OPO staff. Given that a recent analysis from the OPTN contractor included the 799 code in its definition of OOS kidneys, suggesting awareness of this practice, the failure of the OPTN contractor to alert the OPTN’s Membership and Professional Standards Committee (MPSC) or the Data Advisory Committee (DAC) represents inadequate stewardship of the OPTN data, which includes these operational data.^7^ Data entry errors and general concerns about the quality of the data in the OPTN registry that includes operational data is a clear concern that has been highlighted repeatedly, including in the HRSA OPTN modernization effort.^17,18,19,20,21^

These OPO coding practices are likely to significantly underestimate the current scope of OOS placements. For example, in 2023, the inclusion of mOOS kidneys in the overall OOS count increased the prevalence of OOS allocation from 15.5% to 17.7% of all transplants. Failure to adequately audit these data is likely to have adverse repercussions for allocation policy, peer review, and process improvement opportunities within the transplant system. In several instances, we identified OPOs in which all kidneys allocated OOS were coded using the 799 or 898 codes, potentially obscuring the true extent of OOS allocation by these OPOs. For example, in 2023 there were three OPOs that performed a significant amount of OOS transplants (>10) yet miscoded all of these OOS transplants using the 799 code.

The rapid increase in miscoded OOS allocation following the implementation of KAS250 warrants immediate attention to ensure the ability to appropriately monitor the allocation system. In recognition that OOS allocation should only be used as a process of last resort, the OPTN’s MPSC requires OPOs to report every organ placed via OOS allocation with a written justification. It is then the MPSC’s responsibility to review each of these instances, although the number of written notifications submitted each year is not actively tracked (personal communication: OPTN) and these are often templated letters with little specific information that can help the MPSC make any meaningful determination.^22^ From 2017 to 2023, the number of allocation deviations reviewed annually by the MPSC increased almost 7-fold.^22^ This has overwhelmed the capacity to monitor or evaluate allocation deviations: at its 2023 meetings, the MPSC reported reviewing 3,082 allocation deviations among all organs, short of the 3,473 allocation deviations among kidneys alone that we identified in 2023.^22^ In response to this growth, the MPSC decided in January 2024 to limit reviews to allocation deviations “of exceptional concern, along with conducting random samplings of OPOs allocations out-of-sequence”.^22^ In light of these limitations, transplant centers should be required to acknowledge and report when they are accepting a kidney that was allocated OOS to provide redundancy and ensure that there is a complete capture of instances when there is a deviation from the allocation system.

Limitations of this study include an inability to include potential OOS-equivalent entries unless they contained one of our prespecified terms (i.e. expedited, aggressive). For example, on manual review we encountered the free text entry “bypass for targeted list” and many similar variations, which likely reflect OOS allocation but cannot be categorized with certainty. It is therefore likely that this analysis underestimates the true prevalence of miscoded OOS allocation.

In conclusion, we found a marked rise in miscoded OOS allocation – a previously undescribed practice that is inconsistent with OPTN guidelines for documenting OOS placement. This practice now accounts for 12% of OOS transplants and 2% of all deceased donor kidney transplants. These findings highlight the need for stricter oversight of kidney allocation deviations including requirements for both OPOs and transplant centers to acknowledge/report OOS transplants and a renewed commitment to data auditing, training and transparency as well as a robust meaningful review of all OOS allocation, including miscoded OOS, to avoid the misuse of this pathway.

## Acknowledgements

The data reported here have been supplied by the Hennepin Healthcare Research Institute (HHRI) as the contractor for the Scientific Registry of Transplant Recipients (SRTR). The interpretation and reporting of these data are the responsibility of the author(s) and in no way should be seen as an official policy of or interpretation by the SRTR or the U.S. Government.

## Data Availability Statement

The data used for this analysis are available upon request to the SRTR.

## Funding

SAH is supported by NIDDK grant K23DK133729. SM is supported by DK116066, EB032910, DK126739, DK130058 and the Kidney Transplant Collaborative. DC is supported by the NIAID grant R25 AI 147393.

## Disclosures

- Dr. Mohan reported receiving personal fees from Sanofi, Specialist Direct and the Health Services Advisory Group; receiving grants from the National Institutes of Health; serving as past chair for the United Network for Organ Sharing data advisory committee and being the deputy editor of *Kidney International Reports*. Dr. Schold reports receiving consulting fees from Sanofi, eGenesis, and Novartis, as well as grants from the NIH/NIDDK, NIH/NHLBI, Kidney Transplant Collaborative, Department of Defense, and One Legacy Foundation. Dr. Schold also reports serving as the Chair of the Data Advisory Committee at UNOS and a member of the Scientific Research Committee for the SRTR. The rest of the authors do not report any disclosures.

## Abbreviations

OPO: Organ procurement organization
OPTN: Organ Procurement and Transplantation Network
SRTR: Scientific Registry of Transplant Recipients
MPSC: Membership and Professional Standards Committee
OOS: Out-of-sequence
mOOS: Miscoded out-of-sequence
eOOS: Explicit out-of-sequence
KDPI: Kidney donor profile index

## Tables and figures

**Supplemental Figure 1:** Flow diagram of study population

**Supplemental Table 1:** Donor characteristics of deceased-donor kidneys transplanted via in-sequence (IS) allocation, explicit out-of-sequence (eOOS) allocation, and miscoded OOS (mOOS) allocation, 2021-2023

**Supplemental Figure 2:** Relationship between proportion of kidneys allocated via mOOS and count of kidneys allocated via mOOS, 2021-2023. Each dot represents one OPO, and red dots correspond to “outlier OPOs” as defined previously.

**Supplemental Figure 3:** Frequency of explicit and miscoded out-of-sequence allocation at the national level (Panel A) and at the transplant center and year level (Panels B-D), 2021-2023

## Notes

### Author Declarations

The IRB of Columbia University Irving Medical Center gave ethical approval for this work.

