## Supplemental Materials for "Nonstandard coding of deceased-donor kidney out-of-sequence allocation leads to underrecognition of allocation deviations"

**Supplemental Material**

Supplemental Methods

Supplemental Figure 1. Flow diagram of study population

Supplemental Figure 2: Relationship between proportion of kidneys allocated via mOOS and count of kidneys allocated via mOOS, 2021-2023. Each dot represents one OPO, and red dots correspond to “outlier OPOs” as defined previously.

Supplemental Figure 3: Frequency of explicit and miscoded out-of-sequence allocation at the national level (Panel A) and at the transplant center and year level (Panels B-D), 2021-2023. Each bar in Panels B-D represents one transplant center.

Supplemental Table 1. Donor characteristics of deceased-donor kidneys transplanted via in-sequence (IS) allocation, explicit out-of-sequence (eOOS) allocation, and miscoded OOS (mOOS) allocation, 2021-2023

**Supplementary Methods**

*Missing Data*

Among all in sequence, explicit out of sequence, and miscoded out of sequence transplants, 30 donors did not have corresponding entries in the deceased donor file and were therefore missing the following variables: age, gender, race, ethnicity, KDPI, blood type, diabetes status, hypertension status, peak creatinine, cause of death, donation after circulatory death, and hepatitis C status. These 30 donors accounted for 49 transplants, including 12 OOS transplants. The match run date for all 49 transplants was between December 29, 2023, and December 31, 2023. These 49 transplants were included in the overall counts of transplants but excluded from comparison tables (Table 1 and Supplemental Table 1).

17 additional donors were missing race and KDPI data. These 17 donors accounted for 28 transplants, including 1 OOS transplant. These donors’ race was classified as “Other or multiracial,” and their KDPI was excluded from descriptive statistics of KDPI.

**Supplemental Figure 1:** Flow diagram of study population.

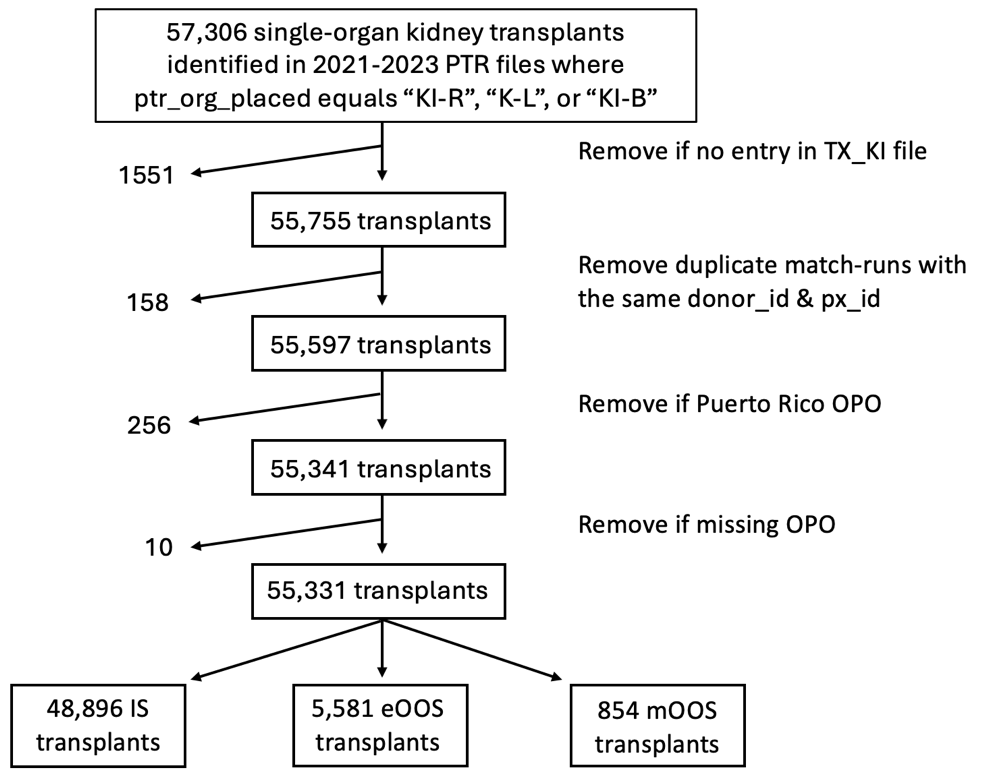

Abbreviations: PTR, potential transplant recipients; OPO, organ procurement organization; IS, in sequence; OOS, out-of-sequence; mOOS, miscoded out-of-sequence; eOOS, explicit out-of-sequence

**Supplemental Figure 2:** Relationship between proportion of kidneys allocated via mOOS and count of kidneys allocated via mOOS, 2021-2023. Each dot represents one OPO, and red dots correspond to “outlier OPOs” as defined previously.

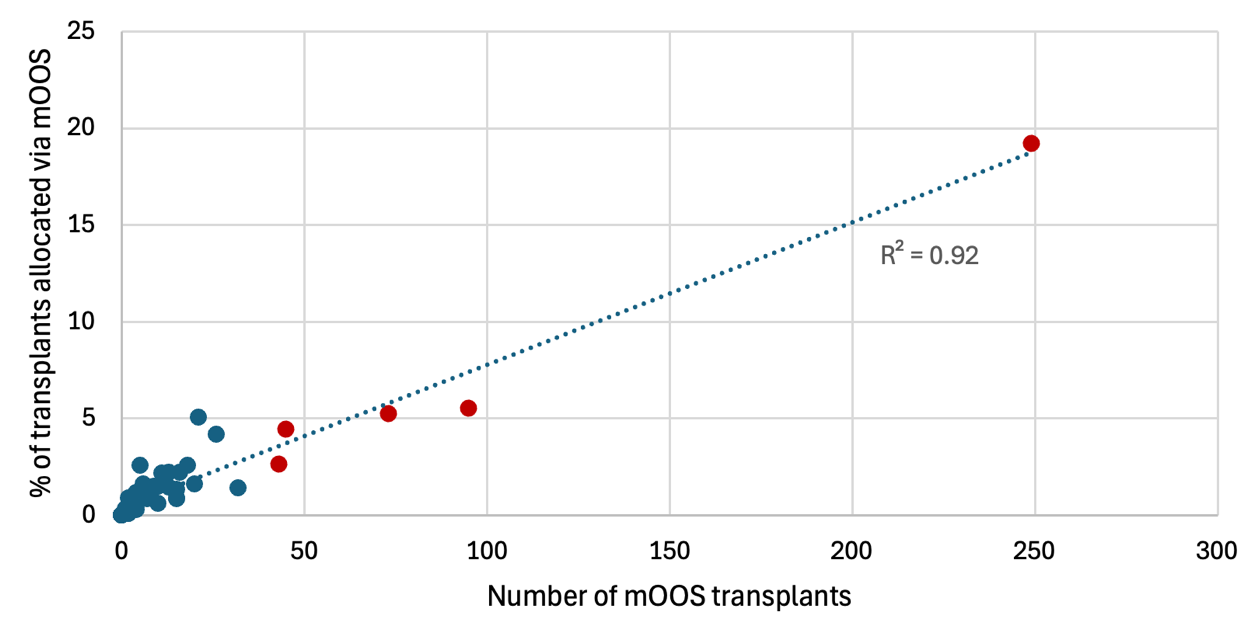

Abbreviations: OOS, out-of-sequence; mOOS, miscoded out-of-sequence; eOOS, explicit out-of-sequence; OPO, organ procurement organization

**Supplemental Figure 3:** Frequency of explicit and miscoded out-of-sequence allocation at the national level (Panel A) and at the transplant center and year level (Panels B-D), 2021-2023. Each bar in Panels B-D represents one transplant center.
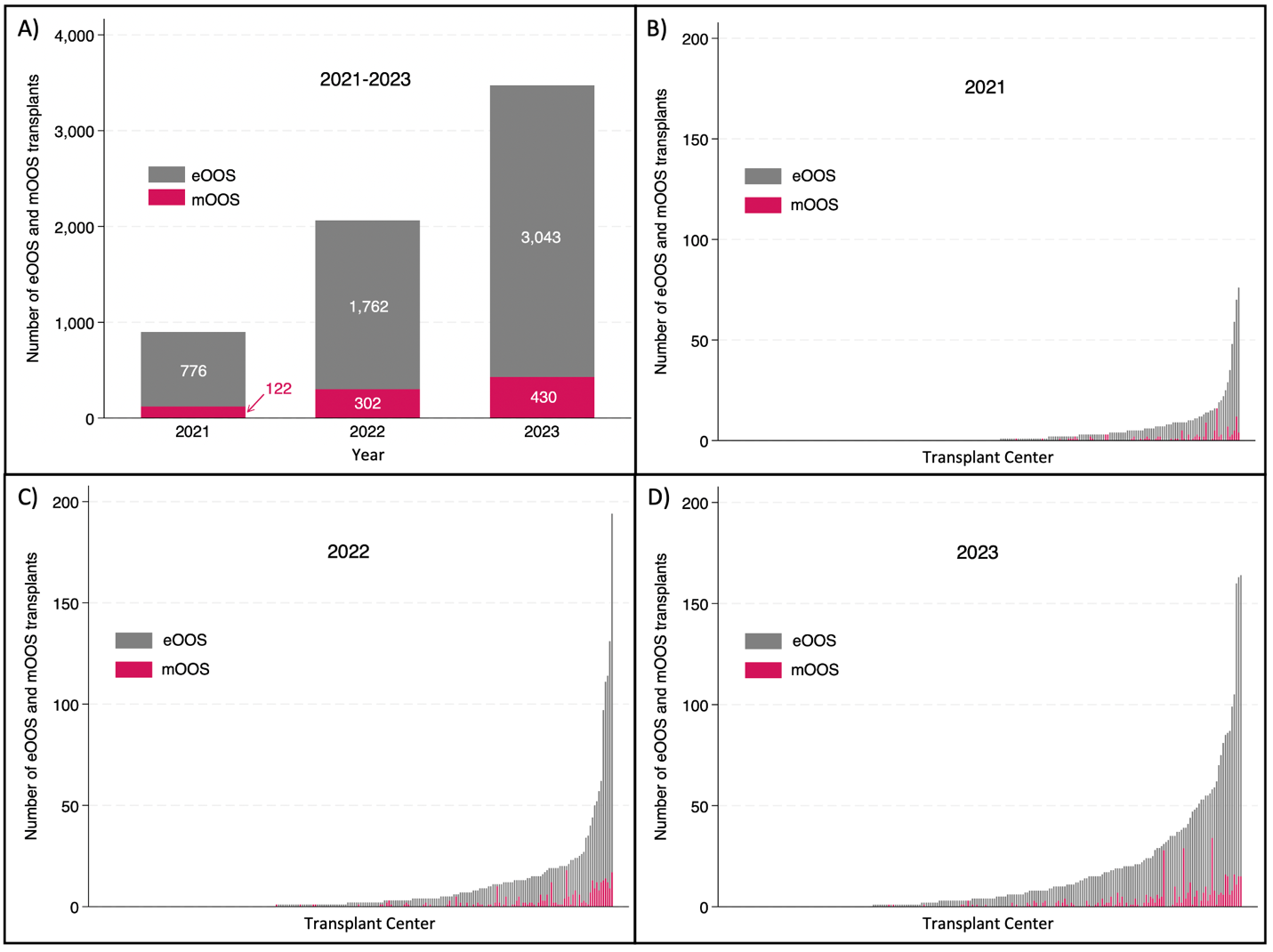
 Abbreviations: OOS, out-of-sequence; mOOS, miscoded out-of-sequence; eOOS, explicit out-of-sequence; OPO, organ procurement organization

**Supplemental Table 1.** Donor characteristics of deceased-donor kidneys transplanted via in-sequence (IS) allocation, explicit out-of-sequence (eOOS) allocation, and miscoded OOS (mOOS) allocation, 2021-2023.

|  | **2021** | | | | **2022** | | | | **2023** | | | |
| --- | --- | --- | --- | --- | --- | --- | --- | --- | --- | --- | --- | --- |
|  | **IS** | **Explicit OOS** | **Miscoded OOS** | **p-value** | **IS** | **Explicit OOS** | **Miscoded OOS** | **p-value** | **IS** | **Explicit OOS** | **Miscoded OOS** | **p-value** |
| **Total** | 16,501 | 776 | 122 |  | 16,223 | 1,762 | 302 |  | 16,172 | 3,043 | 430 |  |
| **Age** | 39 (29-51) | 43 (32-52) | 47 (29-56) | <0.01 | 39 (29-51) | 45 (34-55) | 43 (31-52) | <0.01 | 40 (30-51) | 46 (34-56) | 45 (35-53) | <0.01 |
| **Gender** |  |  |  |  |  |  |  |  |  |  |  |  |
| Female | 5,976 (36%) | 289 (37%) | 42 (34%) | 0.77 | 5,779 (36%) | 644 (37%) | 111 (37%) | 0.69 | 5,796 (36%) | 1,161 (38%) | 153 (36%) | 0.04 |
| Male | 10,525 (64%) | 487 (63%) | 80 (66%) |  | 10,444 (64%) | 1,118 (63%) | 191 (63%) |  | 10,339 (64%) | 1,871 (62%) | 276 (64%) |  |
| **Race** |  |  |  |  |  |  |  |  |  |  |  |  |
| Asian | 390 (2%) | 17 (2%) | <10 (2%) | 0.66 | 397 (2%) | 39 (2%) | 4 (1%) | 0.39 | 418 (3%) | 60 (2%) | <10 (1%) | <0.01 |
| Black | 2,257 (14%) | 103 (13%) | 17 (14%) |  | 2,321 (14%) | 277 (16%) | 48 (16%) |  | 2,213 (14%) | 513 (17%) | 50 (12%) |  |
| White | 13,599 (82%) | 17 (2%) | 102 (84%) |  | 13,249 ( 82%) | 1,412 (80%) | 246 (81%) |  | 13,159 (82%) | 2,416 (80%) | 374 (87%) |  |
| Other or multiracial | 255 (2%) | 639 (82%) | 0 (0%) |  | 256 (2%) | 34 (2%) | 4 (1%) |  | 345 (2%) | 43 (1%) | <10 (0%) |  |
| **Ethnicity** |  |  |  |  |  |  |  |  |  |  |  |  |
| Hispanic | 2,469 (15%) | 121 (16%) | 16 (13%) | 0.75 | 2,491 (15%) | 240 (14%) | 37 (12%) | 0.06 | 2,492 (15%) | 377 (12%) | 35 (8%) | <0.01 |
| Non-Hispanic | 14,032 (85%) | 655 (84%) | 106 (87%) |  | 13,732 (85%) | 1,522 (86%) | 265 (88%) |  | 13,643 (85%) | 2,655 (88%) | 394 (92%) |  |
| **KDPI** |  |  |  |  |  |  |  |  |  |  |  |  |
| Median (IQR) | 42 (22-64) | 54 (30-71) | 49 (27-69) | <0.01 | 40 (20-61) | 55 (34-73) | 48 (29-70) | <0.01 | 38 (19-59) | 54 (33-72) | 53 (33-66) | <0.01 |
| ≥ 85% | 1,154 (7%) | 66 (9%) | 10 (8%) | 0.25 | 966 (6%) | 198 (11%) | 29 (10%) | <0.01 | 845 (5%) | 300 (10%) | 32 (7%) | <0.01 |
| **Blood type** |  |  |  |  |  |  |  |  |  |  |  |  |
| **A** | 6,126 (37%) | 298 (38%) | 40 (33%) | 0.10 | 5,944 (37%) | 606 (34%) | 115 (38%) | <0.01 | 5,911 (37%) | 1,090 (36%) | 185 (43%) | <0.01 |
| **B** | 1,883 (11%) | 80 (10%) | 16 (13%) |  | 1,882 (12%) | 233 (13%) | 35 (12%) |  | 1,956 (12%) | 365 (12%) | 51 (12%) |  |
| **AB** | 571 (3%) | 15 (2%) | <10 (1%) |  | 629 (4%) | 28 (2%) | <10 (2%) |  | 602 (4%) | 45 (1%) | <10 (2%) |  |
| **O** | 7,921 (48%) | 383 (49%) | 65 (53%) |  | 7,768 (48%) | 895 (51%) | 147 (49%) |  | 7,666 (48%) | 1,532 (51%) | 185 (43%) |  |
| **Diabetes** | 1,297 (8%) | 77 (10%) | 11 (9%) | 0.07 | 1316 (8%) | 196 (11%) | 38 (13%) | <0.01 | 1,463 (9%) | 381 (13%) | 50 (12%) | <0.01 |
| **Hypertension** | 4,619 (28%) | 260 (34%) | 37 (30%) | <0.01 | 4,342 (27%) | 663 (38%) | 117 (39%) | <0.01 | 4,583 (28%) | 1,152 (38%) | 162 (38%) | <0.01 |
| **Peak Cr** | 0.9 (0.7-1.4) | 1.0 (0.7-1.9) | 1.0 (0.6-1.7) | <0.01 | 0.9 (0.7-1.4) | 1.1 (0.7-2.1) | 1.1 (0.7-2.1) | <0.01 | 0.9 (0.6-1.4) | 1.1 (0.7-2.0) | 1.0 (0.7-1.7) | <0.01 |
| **Cause of death** |  |  |  |  |  |  |  |  |  |  |  |  |
| Anoxia | 8,022 (49%) | 403 (52%) | 62 (51%) | <0.01 | 8,009 (49%) | 975 (55%) | 163 (54%) | <0.01 | 8,325 (52%) | 1,561 (51%) | 218 (51%) | <0.01 |
| Stroke | 3,423 (21%) | 156 (20%) | 26 (21%) |  | 3,228 (20%) | 355 (20%) | 59 (20%) |  | 3,025 (19%) | 637 (21%) | 103 (24%) |  |
| Head Trauma | 4,519 (27%) | 158 (20%) | 28 (23%) |  | 4,283 (26%) | 327 (19%) | 69 (23%) |  | 4,162 (28%) | 637 (21%) | 81 (19%) |  |
| CNS tumor | 53 (0%) | <10 (1%) | 0 (0%) |  | 50 (0%) | 6 (0%) | 0 (0%) |  | 49 (0%) | 17 (1%) | <10 (1%) |  |
| Other | 484 (3%) | 55 (7%) | 6 (5%) |  | 653 (4%) | 99 (6%) | 11 (4%) |  | 574 (4%) | 180 (6%) | 23 (5%) |  |
| **DCD** | 5,082 (31%) | 335 (43%) | 54 (44%) | <0.01 | 5,102 (31%) | 708 (40%) | 101 (33%) | <0.01 | 5,425 (34%) | 1,407 (46%) | 189 (44%) | <0.01 |
| **Hepatitis C** | 1,661 (10%) | 81 (10%) | 8 (7%) | 0.41 | 1,693 (10%) | 177 (10%) | 29 (10%) | 0.79 | 1,610 (10%) | 300 (10%) | 55 (13%) | 0.15 |

Abbreviations: DCD, Donation after circulatory death; Cr, creatinine, OOS, out-of-sequence; KDPI, kidney donor profile index; CNS, central nervous system
